# Biofortification and Fortification of Wheat Flour: Qualitative analysis for implementation and acceptance

**DOI:** 10.1101/2024.07.31.24311298

**Authors:** Rahima Yasin, Zahra A. Padhani, Mushtaque Mirani, Muhammad Khan Jamali, Mahwish Memon, Sana Khatoon, Riya Rai, Areeba Rahman, Anushka Attaullahjan, Jai K. Das

## Abstract

**Background:** This paper comprehensively investigates various aspects of dietary behaviors relating to the usage of wheat flour and sociocultural preferences embedded within rural communities and aims to bridge health gaps resulting from zinc deficiency by introducing zinc bio-fortified and fortified flour in Pakistan.

**Method:** A household and a market study was conducted in Ghotki and Tando Muhammad Khan districts in Sindh, Pakistan. Various stakeholders involved in the wheat-flour industry including farmers, seed suppliers, owners of atta-chakkis and flour mills, grocers and flour merchants, bakers and hoteliers, consumers, and agronomists were interviewed to gauge their knowledge of bio-fortified and fortified wheat-flour.

**Results:** Wheat-flour is a staple food item in Pakistan, however, agricultural output in Pakistan varies across all provinces. Factors that hinder agricultural productivity include a shortfall of essential resources such as irrigation water, superior quality seeds and fertilizers, and machinery. Farmers use primitive methods of farming as they do not have access to modern technologies, information, or training. Wheat flour market vendors and consumers lack awareness of bio-fortified and fortified wheat flour products and believe the only way to create a customer-base is by ensuring that fortified wheat products are available to all and competitively priced compared to traditional options. Additionally, participants misconstrue the process of fermentation and perceive it as unhealthy. The lack of financial resources and awareness restricts adequate promotion of nutrient-rich food products amongst stakeholders involved in the wheat flour industry. Mass awareness campaigns, education and government incentives could bridge the gaps present and encourage wider adoption of bio-fortified and fortified wheat flour.

**Conclusion:** Policy makers and communities can draw on the recommendations made in this paper to introduce and promote zinc bio-fortified and fortified flour in settings where zinc deficiency is prevalent.

## Background

Most low- or middle-income countries (LMICs) are affected by the double burden of malnutrition (DBM), which refers to the coexistence of undernutrition alongside overweight and obesity. A growing number of the lowest income quartile countries have been reported to have the most severe cases of the double burden of malnutrition (DBM) (1).

Malnutrition is a global health issue with various implications for physical and mental well-being. Promoting access to a diverse and nutritious diet, improving sanitation and hygiene, and providing education on proper nutrition are amongst a few strategies aimed at tackling malnutrition. Micronutrient-related malnutrition, sometimes referred to as “Hidden Hunger,” affects over two billion people and is characterized by deficiencies of vital minerals and vitamins that can lead to several life-threatening health complications (2). Micronutrient deficiency disproportionately impacts children and women of reproductive age groups, especially in LMICs. Crucial micronutrients such as zinc, iron and vitamin A are required for growth and development, deficiencies of which contribute to morbidity in children and adults as well as adverse pregnancy outcomes.

Pakistan ranks 130th out of 195 countries on the Global hunger Index (3). The National Nutritional Survey (NNS) of Pakistan reports that 18.6% of children and 22.1% of women had zinc deficiency. Pregnant women make up 37.5% of those with zinc deficiencies. More than half (53.7%) of the children under five years of age are anemic, with 5.7% of those children suffering from severe anemia. It is projected that 51.5% of children are deficient in vitamin A, 18.6% are deficient in zinc, and 62.7% of children under five are estimated to be deficient in vitamin D (4)

Zinc deficiency is a significant global health issue that afflicts millions of people, particularly in low-resource settings. Essential to several vital metabolic processes, zinc deficiency manifests as a rather nonspecific deficiency, and varies in severity according to age (5). Zinc deficiency among women of reproductive age group (6) and children is a significant public health (7). Consuming foods high in zinc, such as meat, poultry, dairy products, nuts, and legumes, is the primary method of obtaining zinc. Zinc intake by women during their antenatal period reduces the risk of childhood wheeze (8) and the frequency of preterm births (9). Zinc supplementation in children is also linked to a reduced incidence of diarrhea and pneumonia (8).

The second sustainable development goal calls for eradication of hidden hunger and malnutrition (10).

Numerous initiatives (11), such as dietary diversification, food fortification, and supplementation programs have been put forth to address global deficiency of zinc, particularly for at-risk demographics. In Pakistan, food fortification has been introduced which is a cost-effective and sustainable approach aimed at improving the nutritional quality of foods. The technique of adding vitamins and minerals to frequently consumed foods during processing to boost their nutritional value is known as food fortification. It is a safe and a cost-effective strategy for improving diets and preventing and managing micronutrient deficiencies (12).

The agricultural industry holds a pivotal position in shaping Pakistan’s economy, accounting for 24% of its GDP (13). Wheat, a staple crop in Pakistan, is vital for ensuring food security and stability. Cultivated over nine thousand hectares of land, wheat yielded over 27 million tons in 2022-2023, contributing 1.9% to the country’s GDP (14). To address malnutrition and increase dietary zinc intake, different wheat varieties biofortified with zinc have been introduced. These include ‘Chitra’, ‘Zincol 2016’, ‘Akbar 2019’ and ‘Nawab 21’ amongst many others (15). Research has shown that people who consume flour biofortified with zinc have a significantly higher daily dietary uptake of zinc, suggesting that bio-fortification interventions may not only have a positive impact on reducing the risk of dietary zinc deficiency in underserved communities, but also in enhancing crop yield (16).

To address micronutrient deficiencies, the government of Sindh has implemented laws mandating biofortification and fortification of wheat/flour with essential micronutrients. However, in spite of these existing legislative measures, effective implementation remains a challenge.

The objective of this qualitative paper is to assess the existing barriers limiting the implementation of these mandates and to examine the current market dynamics and consumer behaviours within the wheat flour industry in Sindh, Pakistan. The study will focus on gaining insight into the current market practices in the wheat flour industry by examining factors such as production processes, distribution networks, pricing, branding, and marketing campaigns. Simultaneously, the study will delve into consumers’ distinct patterns regarding their wheat flour consumption by assessing their dietary habits, purchasing preferences, sociocultural preferences, awareness of nutritional benefits, and willingness to pay a premium for healthier options. This analysis aims to promote sustainable solutions for biofortifying and fortifying wheat/flour by drawing on recommendations made by our respondents and to subsequently improve acceptance and uptake of biofortified/fortified wheat/flour within zinc-deficient communities, as well as enhance overall public health in the region.

## Materials and methodology

### Study Design

This study has two qualitative components, a market analysis and a household analysis. The intention of the research was to better understand domestic decision-making models, purchasing behaviours and to encourage the use of biofortified and fortified wheat. In the market analysis, we investigated the availability and supply of wheat flour [chakki (formal and informal) and roller] and fortifiable/bio-fortified wheat in different seasons (post-harvest and between harvests).

### Study Setting

With a total population of 207 million in Pakistan (2017), Sindh is the second largest populated province at 47 million people. The study was carried out in two districts of Sindh (i.e., Tando Muhammad Khan (TMK) and Ghotki with a main representation of peri-urban and rural areas and these districts were chosen due to a high prevalence of zinc deficiency with 45.5% and 37% of children under the age of five years having zinc deficiency respectively.

### Study Participants and Recruitment

Study participants were recruited through purposive sampling, based on the study’s aims and objectives. Additionally, snowballing technique was adopted and research participants were asked to assist us in identifying other potential subjects.

We ensured that our sample was ethnically and geographically diverse. Participants/key stakeholders included:

- Farmers
- Distributors and Suppliers
- Millers (formal and informal)
- Sellers
- Naan Shop Owners
- Branded and local bakeries
- Processed food industries
- Restaurants
- Flour Mill Association and Farmer Association
- Representatives from Agriculture and Food Department form provincial and district government
- Community members (male and females)

### Questionnaire Tools

A semi structured interview guide was developed with predetermined open-ended questions after extensive literature search and talking to experts. The guide was created in English language and then translated into the locally spoken Urdu and Sindhi language.

The interview guide facilitated the organization, analysis, and synthesis of documentation on factors influencing the wheat supply chain and purchasing behaviors. In-depth interviews (IDIs) and focal group discussions (FGDs) were employed to investigate various aspects, including the current state of wheat cycle, biofortification, fortification as well as political, sociocultural, financial, and infrastructural barriers related to wheat biofortification and wheat flour fortification. The questions captured self-reported food-related behaviors and experiences related to growing challenges in obtaining food due to limited resources. Additionally, the questionnaire included questions pertaining to the cultivation and storage practices of wheat flour, scale of consumption of wheat flour, preference for different brands of wheat flour, market mechanism of wheat flour and business practices, as well as the scope of knowledge about fermentation, biofortification and fortification practices.

### Data Collection and Analysis

The data collection for household and market analysis was conducted between November 25^th^, 2021, to February 17^th^, 2022. The field staff visited households and markets during working hours to maximize the likelihood of finding participants. All interviews were conducted in private spaces of offices and homes of the participants to ensure that confidentiality and integrity of the interviews was maintained throughout. Interviews were conducted until a point of saturation was reached in research, suggesting no new or significant information was emerging from additional interviews. Informed written consent was taken from all the participants of the study. All participants were informed about the right to refuse or withdraw at any time from the study without prejudice.

Data was collected using IDIs and FGDs. The conversations took 40-60 minutes and were audio-recorded with the consent of the participants. All IDIs and FGDs were transcribed verbatim in either Sindhi or Urdu. This was then translated to English and checked by another researcher.

NVivo software was used to manage and code interviews after data collection, and thematic analysis was carried out. We applied the thematic approach developed by Braun and Clarke (17) which included the seven steps of transcription/translation, reading and familiarization, coding, searching for themes, reviewing themes, defining, and naming themes and finalizing the analysis. RY and AR read and reread through each transcript to gain familiarity with the content and context. Both researchers individually performed inductive coding on the first two interviews with each stakeholder to create a coding scheme, which was then analyzed, followed by coding of the remaining transcripts. All responses were coded to relevant nodes, which were later categorized into hierarchy of sub-nodes. Latent content analysis was used to analyze the data. Emerging themes and subthemes that would aid in addressing our research objectives were selected from the tree nodes after the entire dataset had been coded. Each translation was reviewed to make sure all relevant data had been recorded and correctly identified, categorized, and interpreted. Critical findings were conveyed through the inclusion of pertinent quotations that were extracted from the interviews. The anonymity of participants was maintained throughout.

### Ethics Statement

The study design, sampling strategy, instruments and analytical plans were reviewed and approved by the AKU Ethical Review Committee (ERC) and the National Bioethics Committee (NBC), Pakistan. Confidentiality of all collected data was assigned high priority at each stage of data handling. Informed verbal and written consent was obtained from all participants of the study. All participants were informed about the right to refuse or withdraw at any time from the survey without prejudice. Respondents were free to stop interviews at any time or skip any questions they did not want to answer. They had the right to ask questions at any point before, during or after the interview.

## Results

We conducted a total of 60 interviews from Ghotki and TMK collectively, of which 18 were IDIs and 42 were FGDs (Table 1). Of 222 participants interviewed, 195 were males and 27 were females (Table 1). The following themes emerged from our research: wheat farming in Sindh, consumer practices, sociocultural preferences, existing perceptions of health, branding, packaging and quality, affordability, challenges and disparities in the flour industry: stakeholders’ perspectives, and solutions (figure 1).

**Table 1:**
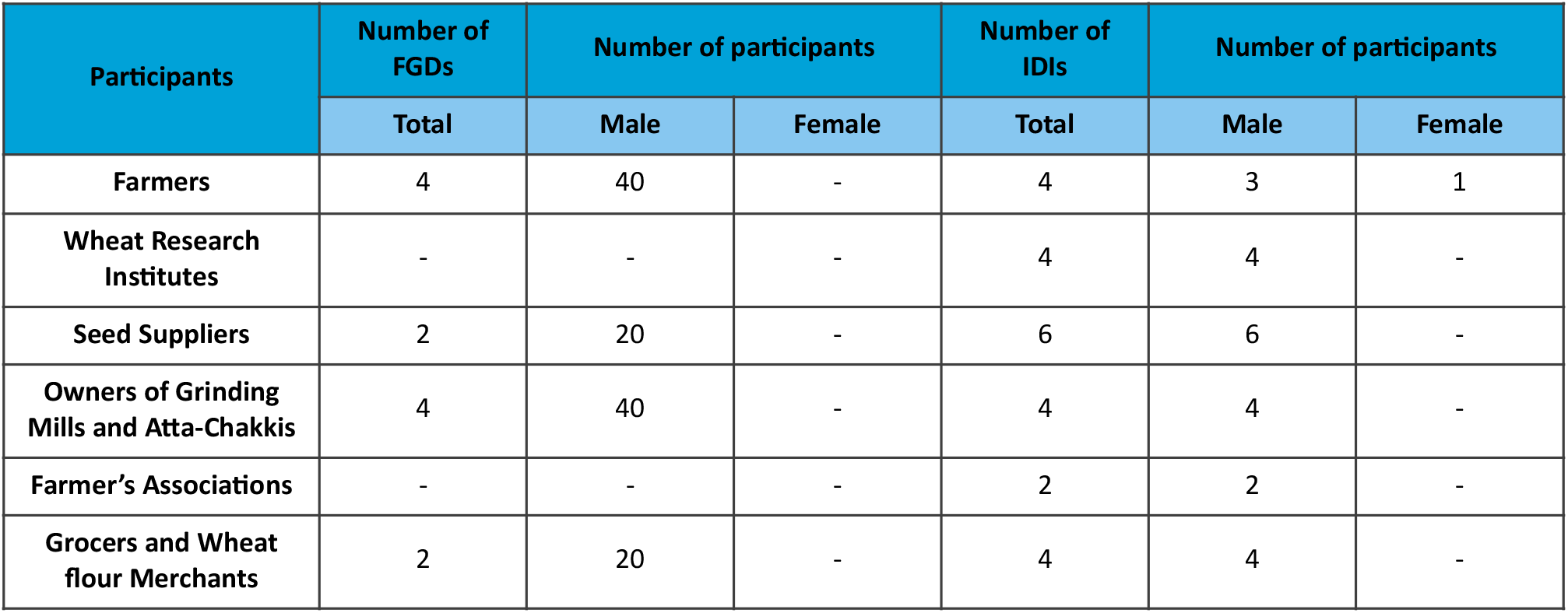
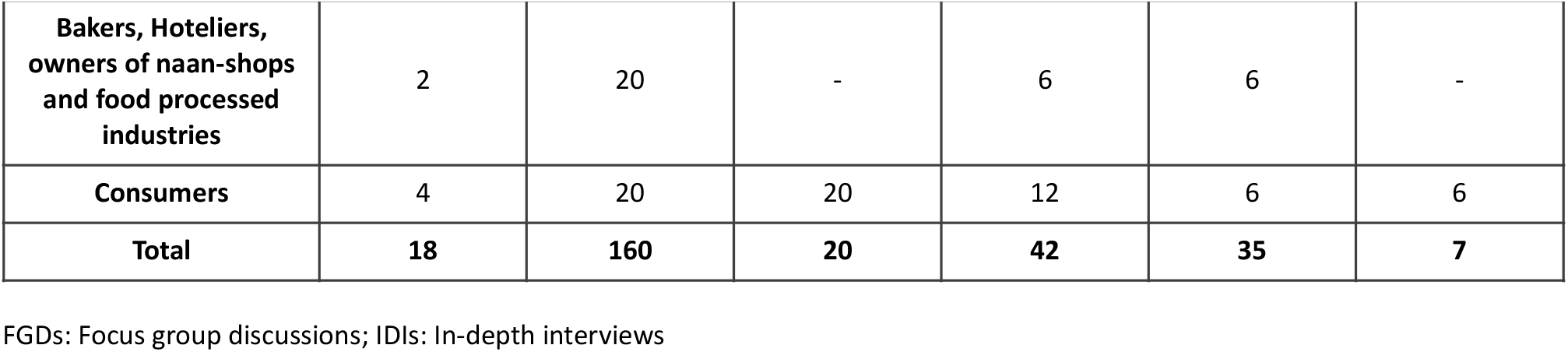
Total number of participants.

| Participants | Number of FGDs | Number of participants |  | Number of IDIs | Number of participants |  |
| --- | --- | --- | --- | --- | --- | --- |
|  | Total | Male | Female | Total | Male | Female |
| Farmers | 4 | 40 | - | 4 | 3 | 1 |
| Wheat Research Institutes | - | - | - | 4 | 4 | - |
| Seed Suppliers | 2 | 20 | - | 6 | 6 | - |
| Owners of Grinding Mills and Atta-Chakkis | 4 | 40 | - | 4 | 4 | - |
| Farmer's Associations | - | - | - | 2 | 2 | - |
| Grocers and Wheat flour Merchants | 2 | 20 | - | 4 | 4 | - |
|  | Total | Male | Female | Total | Male | Female |
| Bakers, Hoteliers, owners of naan-shops and food processed industries | 2 | 20 | - | 6 | 6 | - |
| Consumers | 4 | 20 | 20 | 12 | 6 | 6 |
| Total | 18 | 160 | 20 | 42 | 35 | 7 |

FGDs: Focus group discussions; IDIs: In-depth interviews

**Figure 1:**
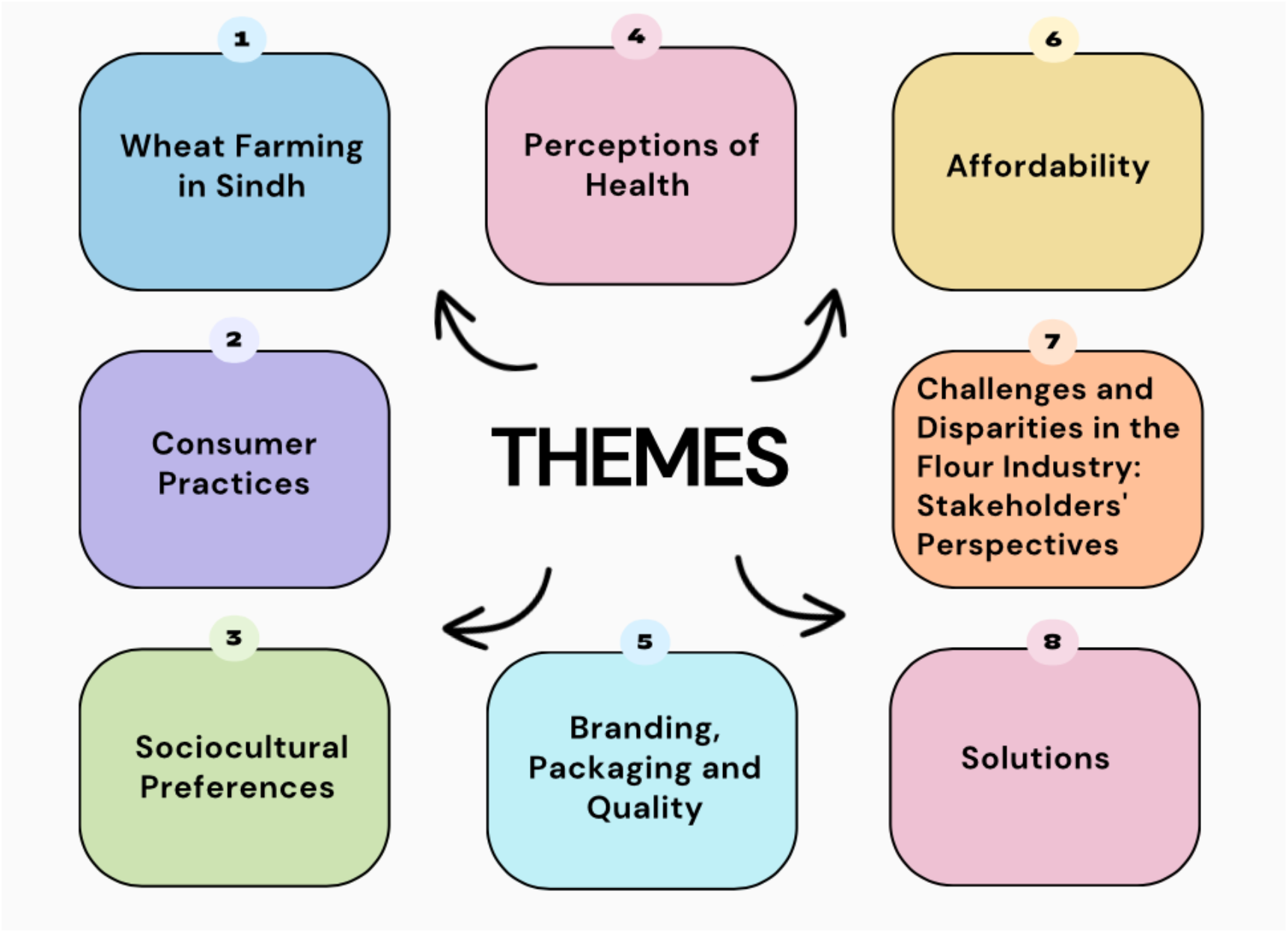
Themes.

### Wheat Farming in Sindh

Sindh’s agricultural plains are significantly less fertile, with less irrigation water than Punjab province in Pakistan. In addition to a shortage of irrigation water, late cultivation of wheat, insufficient supply of phosphate fertiliser, weather variations, absence of superior or certified seeds, and less-than-ideal land conditions, which all contribute to low yield.

Farmers plant wheat not just for human consumption, but also to feed their livestock in case of shortage of green-grass. However, wheat cultivation has decreased over time, as farmers choose cash crops over wheat due to its high profitability. They prefer to cultivate and utilise their own seeds, but it is also common practice in their community to purchase seeds from local seed providers, merchants, and other farmers in times of scarcity. Landlords, particularly those with established operations, store their own seeds for the forthcoming cultivation period, whereas smaller-scale landlords procure them from local seed companies.

Farmers prefer variety of seeds that not only provide a high yield but are also inexpensive, easy to cultivate, resistant to illnesses, and require little water. One specific zinc bio-fortified seed variety, ‘Akbar-19’, is popular in Ghotki because to its high yield; yet, locals are unaware of its nutritional advantages. Moisture content is a crucial factor to consider while storing seeds since it makes them more susceptible to different diseases such as rust, and insect infestations. Once the wheat has been cultivated, farmers let the seeds dry in the sun for a few days, then filter them to remove any impurities, husk, and weak grains. They then add “neem” leaves and sometimes even red chilies and store them in aluminium containers.

“*Actually, wheat is more profitable than other crops as we can utilize it for two purposes - consuming ourselves and selling it in the market. It*’*s a biannual crop, and we can cultivate cotton crops to feed our animals or paddy crops in the other seasons on the same land*.*”*

### Consumer Practices

A significant number of consumers in rural regions grow and harvest their own wheat, and personally oversee the grinding of wheat at atta-chakkis to ensure that no chemicals are introduced to the product. This practice reflects a deep-rooted trust and preference for self-produced food over externally purchased items.

In case consumers prematurely deplete their flour reserves, they purchase flour from utility stores at subsidised rates or preferably from atta-chakkis or grinding mills, either on cash or credit, depending on their financial liquidity. In case of surplus inventory, wheat flour is sold in the market. Some participants also voiced their suspicion of millers mixing sand and mud in the flour.

Wheat is one of the most common staple and households usually knead the dough at mealtimes to keep the bread or roti fresh. They make an assortment of wheat flour-based items at home, including different types of breads, porridge, and traditional desserts, and purchase different wheat flour-based products like potato pastries, puff pastries, biscuits, cakes, and specialty treats from their local market. They also add flour to curries to give the curries a thicker texture.

“*We use wheat flour after wheat crop is harvested, from the beginning of March to August. After this, our wheat stock finishes, and we buy wheat flour from retail shops until rice crop is harvested. We usually buy special wheat flour from the market*.*”*

### Sociocultural Preferences

In TMK, wheat vendors mentioned that customers who belonged to urban areas, had a higher demand for special flour due to the colour, texture and taste of its bread, and the fact that the dough was simply easier to kneed. This preference was followed by whole-wheat flour, refined flour and semolina. Participants also mentioned that there were no flour mills in TMK as their region was more suited for the cultivation of rice crop.

“*Due to water scarcity in wheat cultivation season and the quality of agricultural land, this area is ideal for rice crop cultivation, but not for wheat flour cultivation. The production of wheat is very low here, that*’*s why there is no any flour mill at TMK*.*”*

Many baked goods, such as breads, utilise fermented flour as it contributes to improved texture, flavour, and volume. However, due to misconceptions about health, consumers only sporadically consume fermented flour-based goods at home, such as pizza, tandoori breads, and cakes. They utilise curd and yeast to ferment flour, a fermentation method that has been taught to them by their ancestors, and this practice of traditional food preparation methods that has been passed down from generation to generation emphasises the importance of cultural heritage.

As opposed to wheat flour consumers at homes, restauranteurs, bakers and naan-shop owners consistently use fermented flour to prepare a variety of breads or rotis and are acquainted with the concept of fermentation. This helps vendors accommodate clients who have a predilection for leavened white breads.

“*We mix refined flour with whole-wheat flour, this gives the chapatis the white color. You must have noticed that rotis and chapatis made at home from whole wheat flour are brown in colour and look untidy. In hotels, we mix in special flour, which gives chapatis a pleasing white colour*.*”*

### Perceptions of Health

Most families prefer using whole-wheat flour due to its bran and fiber content, and the texture and colour it gives to the bread or *roti*. They believe that refined *atta* yields breads and *rotis* which are harder to chew, more difficult to digest and which eventually lead to constipation. Flour consumers at home prioritise unprocessed flour due to perceived health benefits.

“*People prefer whole wheat flour. We too believe it is the best amongst all and is healthier. As compared to the past, nowadays the use of pesticides and artificial fertilisers to increase yield has reduced the quality of wheat. But whole wheat flour is still more nutritious and good for our health*.*”*

Additionally, consumers also expressed concerns of poor digestion associated with fermented flour and deemed it unhealthy.

The small number of respondents who do consume bio-fortified and fortified flour are unaware of its nutritional advantages and simply consume it because they believe it is of higher quality, and is conveniently accessible in department stores.

According to some respondents, the sole motivation for eating flour is to simply alleviate their hunger and to provide basic sustenance to themselves and their families.

### Branding, Packaging and Quality

Since most participants in Ghotki and TMK districts hail from economically-disadvantaged backgrounds and lack literacy, they are unable to read labels or nutritional information on packaging, and therefore sometimes perceive attractive packaging and logos as deceptive. However, a few maintained that branded flour is healthier and of higher quality.

“*Customers are satisfied in simple packaging too, they just need good quality. If we do not provide good quality to them, they will definitely get a bad impression and won’t come back to us*.*”*

Some respondents claim to be selective about brands they prefer and feel the flour’s quality varies depending on the mill it is purchased from, as some sell superior flour with a high gluten content, while others sell impure flour unfit for use.

As majority of local hotels and naan-shops in the Ghotki and TMK community operate on a small scale and lack the means for proper branding, they rely solely on recommendations from their customers for marketing. It is only the well-established businesses that prioritise the importance of effective branding and invest significantly to harness its full potential.

Special flour comes in elegant packaging, with logos exclusive to the mills that manufacture it. This makes it easier for customers to identify the flour and ascertain its source, allowing them to assess its quality. Alternatively, atta-chakkis sell basic or fine flour in simple plastic bags and unmarked bags.

### Affordability

When purchasing flour, consumers are more influenced by the cost of the flour than by its nutritional value. Respondents, particularly those from the Ghotki area, stated that they purchase the cheapest flour available and that if the price of their favoured brand rises, they switch to a cheaper alternative. Participants in the TMK region, on the other hand, are more inclined towards purchasing flour variants that are both fresh and high in fibre.

Consumers alleged that local merchants and dealers were manipulating prices and charging exorbitant prices for wheat flour, despite the government providing them at regulated rates. Consumers requested a check and balance on flour pricing and for the food department officials to take strong action against those who unlawfully inflated the prices of flour.

Most community members who were familiar with biofortified and fortified flour voiced reservations about its perceived high costs. A paucity in available options is another element influencing the sale of biofortified and fortified flour. In rural areas, buyers have restricted alternatives because there is only one brand of fortified flour available which is more expensive than plain flour. Vendors believe the high costs of fortified flour creates a barrier in its sale.

All consumers agreed to switching to bio-fortified wheat varieties and fortified flour only if it indeed was a healthier option, and the government provided it either for free initially on trial-basis, or at relatively cheaper rates.

### Challenges and Disparities in the Flour Industry: Stakeholders’ Perspectives

Stakeholders who use flour commercially allege that the prices of fertilisers and wheat flour proposed and sanctioned by the government are never implemented and are unreasonably hiked by black-market dealers. Farmers purchase agri-inputs on credit due to poverty and often sell their wheat back to merchants from whom they purchased agri-inputs to pay off their debt. They lack the right machinery and efficient tools and must put in manual labor while harvesting wheat. Small-scale farmers are also unable to procure wheat-grains, fertilizers and pesticides in time for cultivation. Many farmers lack training and expertise when it comes to cultivating a crop and rely on primitive knowledge passed down from their forefathers. The absence of facilities and technology in agriculture poses significant challenges for farmers and limits their productivity and eventually affects livelihoods. Respondents pointed out the absence of active government involvement in the agricultural sector, especially in the province of Sindh.

Though the government offers comparatively better rates for wheat grain than trade commission agents, the government officials often treat poor farmers who are simply trying to make ends meet differently from wealthier farmers. This discrimination takes the form of delayed payments or refusal of provision of government-approved stamped wheat bags. The impoverished farmers are forced to sell their goods to local trade commission brokers as a result, who frequently take advantage of them and put them in a deficit.

“*Farmers in Sindh live hand-to-mouth. We don*’*t have enough resources to get new machines and technology. Government and banks don*’*t support us, don*’*t offer interest-free or easy-financing services, nor do we get any subsidies when it comes to agricultural tools and machines*.*”*

Chakki-owners in rural areas are not registered with the government as the participants claimed the process of registration was complicated and involved various hurdles that only influential businessmen could get past. They don’t follow a financial model and make transactions via cash, credit and maintain a basic register to summarise their sales. They find it difficult to keep operational costs of their businesses low. They are unable to afford electricity as it is very expensive and load-shedding is common, and therefore run their machinery on diesel generators. At the end of the month, chakkis in rural areas are only able to make a profit of five-to-ten thousand rupees or just around thirty dollars. In comparison, mill-owners in urban areas are registered with the government, and maintain a ledger cash book, balance sheets and registers and relatively financially secure.

If the government raises prices or there is a scarcity of flour, mill owners are compelled to buy grain at high rates on the open market, having no choice but to raise their own prices. As a result, they sell the flour in the market at inflated rates.

Wheat flour merchants and vendors source flour from flour-mills, grinding mills, and wholesalers in bulk quantity at lower costs in an effort to price it reasonably for their clientele. However, they allege that mill-owners do not follow through on their reduction of rates from the government and instead sell flour at high costs. Flour merchants are concerned that their businesses would become unbearably costly to run owing to the escalating prices, as well as the steep rent and utility charges. For the sake of their customers, some participants seek to keep their flour prices reasonable, even if it calls for a reduction in their own profit margin.

“*We are bearing high rents of our shops, and utility expenses are increasing day by day. If we cannot earn our livelihood, how can we run this business?”*

### Solutions

In low-resource settings, limited funding, lack of technologies, inadequate infrastructure, food insecurity and low literacy rates impede progress and growth in the agricultural sector. It is, therefore, necessary to bridge these gaps to optimize agriculture productivity. One of the vital steps towards achieving this is augmenting government support. Stakeholders demanded the government play a more active role by ensuring the implementation of controlled and regularized rates of fertilizers and flour in the market, to prohibit black market dealers from meddling with the fixed costs set by the government and to timely tackle and prevent the artificially created shortage of fertilizers, wheat-grain and wheat flour. It is imperative for the government officials to not discriminate between small-scale and large-scale farmers and to make government-approved stamped wheat bags available to all. Additionally, providing farming equipment, machineries including tractors, different agricultural inputs and fresh grains to farmers, premixes for fortification to chakki-owners and fortified wheat flour to consumers at subsidized costs could also encourage stakeholders to try out the new flour variant. Making fortified wheat flour instead of regular flour a part food-relief programs could also encourage consumers to try the fortified flour variant. Challenges such as frequent power outages, inadequate infrastructure and lack of security need to be addressed. Participants proposed the practice of cultivating new varieties of wheat mandatory for landlords and farmers.

Research scientists emphasized strengthening government support by establishing laboratories for seed-testing which could, most importantly, be accessible in rural areas as well as be affordable to small-scale farmers. The laboratories would evaluate the seeds for their germination capacity, moisture content and provide an overall assessment of the seed’s planting value.

Distribution of literature amongst farmers on methods to keep the crop disease-free and optimize the crop’s yield could be constructive. Community-based awareness and educational programs disseminating key messages about the yield, production, nutritional value and health benefits of biofortified and fortified flour could act as a catalyst for its success. Advertising and creating a demand for biofortified and fortified flour in the market through social media, television, FM radio, brochures and pamphlets could be used as an effective marketing strategy.

*“Due to media exposure, people are now more conscious and concerned about their own and their family’s health. If people start demanding fortified flour, then it will become commonly available. There is room for improvement in the market*.*”*

Participants suggested initiating the promotion of fortified flour from government hospitals, where doctors could recommend using fortified flour to their patients. Agricultural schools, especially in low-resource countries like Pakistan, could also play a crucial role in promoting sustainable farming practices and techniques, and subsequently improving livelihoods in rural areas.

Participants called for awareness programs and better advertisements of bio-fortified varieties and fortified wheat flour so that they could offer their families healthier and more nutritious meals. All interviewed stakeholders had similar perspectives and offered valuable recommendations which could potentially be implemented to optimize the use of fortified flour as part of a regular diet.

*“The world is changing day-by-day, and looking into the health of people, it is necessary to have nutritional options for them, there is a scope of business of products made from fortified wheat flour, but we need to create a market for it*.*”*

## Discussion

The findings from our analysis suggest shared priorities amongst different stakeholders involved in the wheat flour industry. From agri-input sellers to consumers, all stakeholders stressed the importance of the government’s support, and to make fortified flour available and affordable to all, and not just to the rich.

The consensus view seems to be that business owners and whole-sellers selling wheat struggle due to minimal profit margins, as there is no price-control mechanism in the wheat flour market system. The only advantage they have for harvesting wheat crops is that it is a biannual crop, and at the end of the season, they can stock enough for their own consumption. Food department authorities need to take strict actions against those who illegally raise the flour prices and maintain a check and balance of the prices of the flour. It is essential to strike a balance between the interests of rural farmers and urban consumers to guarantee that farmers are fairly compensated for their produce and that consumers may obtain reasonably priced, superior-quality food.

Only a few stakeholders associated with these industries are vaguely familiar with the idea of fortification, having utilized the practice of fortifying flour at their mills While several Ghotki participants were grateful that UKAID and USAID had previously provided them with fortification feeders, premixes, and training programmes on how to fortify flour under the Food Fortification Programme (FFP), the programme was regarded inefficient because of a lack of awareness.

Consistency in marketing strategies and branding across different social media platforms is imperative. It has been a consensus that proper advertisement to create a market for food products made with fortified-flour and awareness of the fortification process through different platforms as well as through health facilities are vital towards making fortified flour popular amongst the public. And while some stakeholders in the market were eager to offer these nutritious options to their customer-base, they were concerned about its niche in the market and the lack of awareness which could potentially result in insufficient demand for fortified food products in the market. Since respondents also misconstrue the addition of chemicals in the flour as a harmful practice, spreading awareness about fortification and addressing misconceptions related to the addition of chemicals in flour is crucial for promoting the adoption of this nutrition-enhancing practice and dispelling any myths. Overcoming these obstacles may lead to a wider range of consumers using fortified flour, which would improve nutritional outcomes within communities.

About 23 percent of Pakistan’s GDP comes from the agricultural sector, which also employs 37.4 percent of the working force. With a land area of 30.5 million hectares, agriculture accounts for over 47% of the country’s total land area, which is greater than the average of 38%. Wheat is a major staple crop in Pakistan and accounts for 37% of total crop area (18).

Agriculture in low-resource settings faces unique challenges, including restricted access to technology, machinery, agricultural inputs and infrastructure. The main obstacles to the adoption of sustainable agricultural practices are knowledge gaps, inadequate extension services, as well as lack of global, national, regional, and local agricultural policies (19). However, with the right strategies, techniques, and policy reforms, agricultural productivity and food security can be improved.

Affordable and locally available technologies, such as hand tools, simple irrigation systems, and pest control methods that do not require expensive machinery or chemicals should be accessible to all farmers. A study by Yadava suggested that farmers would be more likely to quickly adopt nutritionally superior cultivars if seeds and other inputs were available at a reduced cost. Similar to the Brix meter used to assess the sugar content in sugarcane and sweetcorn, the study recommended creating a tool or piece of equipment that would be easy to use and accurately measure the quality parameters of the crop (20).

The significance of biofortification and fortification strategies lies in their ability to enhance the nutritional quality of staple foods. Consumption of biofortified wheat flour has shown to significantly improve plasma zinc levels (21). Another study demonstrates that zinc-biofortified flour resulted in an increase in intakes of zinc and iron (22). Food biofortification and fortification is frequently touted as one of the most cost-effective public health measures (23). Many countries have regulatory bodies or agencies in place to mandate and monitor food fortification programs. These entities provide standards, guidelines, and regulations for fortification levels, to ensure safety and efficacy (24). This intervention plays an imperative role in tackling widespread nutrient deficiencies and improving public health outcomes. Additionally, biofortification and fortification of a staple crop such as wheat flour can serve as a cost-effective strategy in populations that heavily rely on wheat flour as a dietary source, and can be easily made accessible to a larger population.

## Conclusion

Agriculture in Pakistan can flourish with the right blend of knowledge, resources, skills and community outreach. Biofortification of wheat-grains and fortification of wheat flour with zinc can be used as an effective strategy aimed at addressing zinc deficiency and subsequently improve public health in settings where access to a nutrient-rich diet is limited. Addressing these issues requires a multifaceted approach that considers the regional context, cultural norms, and economic constraints while striving to improve the nutritional quality of food, especially in rural communities.

## Data Availability

data available on request

## Author Contributions

M.Mirani, M.K.J., M.Memon, S.K. R.R. carried out the interviews and translated the interviews. R.Y. and A.R. reviewed and cross-checked the translations. R.Y. and Z.A.P. carried out the qualitative analysis. R.Y. drafted the manuscript. A.A. and J.K.D. provided supervision for each step, conceptualization and contributed to the critical revision of the manuscript.

## Data availability

Data available on request.

## References

1. Popkin BM, Corvalan C, Grummer-Strawn LM. Dynamics of the double burden of malnutrition and the changing nutrition reality. Lancet. 2020;395(10217):65–74.

2. Global Hunger Index 2014. https://www.ifpri.org/sites/default/files/ghi/2014/index.html. Accessed on 9th November 2023.

3. Pakistan Global Health Index 2021. https://www.ghsindex.org/country/pakistan/. Accessed on 9th November 2023.

4. Pakistan National Nutrition Survey 2021. https://www.unicef.org/pakistan/media/1951/file/Final Key Findings Report 2019.pdf. Accessed on 9th November 2023.

5. Shah D, Sachdev HS, Gera T, De-Regil LM, Peña-Rosas JP. Fortification of staple foods with zinc for improving zinc status and other health outcomes in the general population. Cochrane Database Syst Rev. 2016;2016(6):Cd010697.

6. Chandyo RK, Strand TA, Mathisen M, Ulak M, Adhikari RK, Bolann BJ, et al. Zinc deficiency is common among healthy women of reproductive age in Bhaktapur, Nepal. J Nutr. 2009;139(3):594–7.

7. Berhe K, Gebrearegay F, Gebremariam H. Prevalence and associated factors of zinc deficiency among pregnant women and children in Ethiopia: a systematic review and meta-analysis. BMC Public Health. 2019;19(1):1663.

8. Li J, Cao D, Huang Y, Chen B, Chen Z, Wang R, et al. Zinc Intakes and Health Outcomes: An Umbrella Review. Front Nutr. 2022;9:798078.

9. Chaffee BW, King JC. Effect of Zinc Supplementation on Pregnancy and Infant Outcomes: A Systematic Review. Paediatric and Perinatal Epidemiology. 2012;26(1):118–37.

10. UNICEF and the Sustainable Development Goals. https://www.unicef.org/sustainable-development-goals. Accessed on 9th November 2023.

11. Roohani N, Hurrell R, Kelishadi R, Schulin R. Zinc and its importance for human health: An integrative review. J Res Med Sci. 2013;18(2):144–57.

12. Olson R, Gavin-Smith B, Ferraboschi C, Kraemer K. Food Fortification: The Advantages, Disadvantages and Lessons from Sight and Life Programs. Nutrients. 2021;13(4).

13. Pakistan Bureau of Statistics: Agriculture 2023. https://www.pbs.gov.pk/content/agriculture-statistics.

14. Finance Division. Government of Pakistan. Pakistan Economic Survey 2022-23. https://www.finance.gov.pk/survey/chapters_23/02_Agriculture.pdf.

15. Wani SH, Gaikwad K, Razzaq A, Samantara K, Kumar M, Govindan V. Improving Zinc and Iron Biofortification in Wheat through Genomics Approaches. Mol Biol Rep. 2022;49(8):8007–23.

16. Rehman A, Farooq M, Ullah A, Nadeem F, Im S, Park S, et al. Agronomic Biofortification of Zinc in Pakistan: Status, Benefits, and Constraints. Frontiers in Sustainable Food Systems. 2020:591722.

17. Clarke V, Braun VJSqr. Successful qualitative research: A practical guide for beginners. 2013:1–400.

18. FAO: Pakistan at a Glance. https://www.fao.org/pakistan/our-office/pakistan-at-a-glance/en/. Accessed on 9th November 2023.

19. Rehman A, Farooq M. Chapter 18 - Challenges, constraints, and opportunities in sustainable agriculture and environment. In: Farooq M, Gogoi N, Pisante M, editors. Sustainable Agriculture and the Environment: Academic Press; 2023. p. 487–501.

20. Yadava DK, Hossain F, Mohapatra T. Nutritional security through crop biofortification in India: Status & future prospects. Indian J Med Res. 2018;148(5):621–31.

21. Lowe NM, Zaman M, Khan MJ, Brazier AKM, Shahzad B, Ullah U, et al. Biofortified Wheat Increases Dietary Zinc Intake: A Randomised Controlled Efficacy Study of Zincol-2016 in Rural Pakistan. Front Nutr. 2021;8:809783.

22. Gupta S, Zaman M, Fatima S, Shahzad B, Brazier AKM, Moran VH, et al. The Impact of Consuming Zinc-Biofortified Wheat Flour on Haematological Indices of Zinc and Iron Status in Adolescent Girls in Rural Pakistan: A Cluster-Randomised, Double-Blind, Controlled Effectiveness Trial. Nutrients. 2022;14(8).

23. Ohly H, Broadley MR, Joy EJM, Khan MJ, McArdle H, Zaman M, et al. The BiZiFED project: Biofortified zinc flour to eliminate deficiency in Pakistan. Nutr Bull. 2019;44(1):60–4.

24. Hennessy Á, Walton J, Flynn A. The impact of voluntary food fortification on micronutrient intakes and status in European countries: a review. Proc Nutr Soc. 2013 Nov;72(4):433–40. doi: 10.1017/S002966511300339X. Epub 2013 Sep 11. PMID: 24020749.

